# Clinicopathological, surgical management and early postoperative outcome among obstructive jaundice inpatients at Kilimanjaro Christian Medical Centre from March 2020 to March 2023

**DOI:** 10.1101/2023.12.05.23299491

**Authors:** Mkunde Ramadhani Chambo, Elmes Venant, David Msuya, Samwel Chugulu

**Affiliations:** General surgeon resident, KCMC Hospital, P.O.Box 3010, Moshi; Tutor Kilimanjaro College of Health and Allied Sciences (KICHAS), P.O.Box 565, Moshi; General surgeon, KCMC Hospital, P.O.Box 3010, Moshi; Head General Surgery department, KCMC Hospital, P.O.Box 3010, Moshi; Consultant Cardiothoracic Surgeon, KCMC Hospital, P.O.Box 3010, Moshi

## Abstract

Obstructive jaundice is a surgical condition that affects all age groups mostly to the above 18 individuals. This condition can lead to high morbidity and mortality if not early diagnosed and treated. It remains the common cause of surgical consultations globally accounting for 8-10/100,000 and 242/1000 annually in China and Saudi Arabia respectively. Clinicopathological and radiological investigations are the key to the disease diagnosis. Cholelithiasis and cancer head of pancreas are common causes of obstructive jaundice. Although the diagnosis and treatments of obstructive jaundice is advancing; some patients get poor treatment outcome such as SSI, peritonitis and long hospital stay with some of them dying.

To determine the clinicopathological, surgical management and early postoperative outcome among obstructive jaundice inpatients at KCMC from March 2020 to March 2023.

A hospital-based cross-sectional study was conducted using secondary data from hospital records, EHMS system, surgery admission and theater registry books. All files of patients aged 18 years and above from these registry books with a diagnosis of obstructive jaundice from March 2020 to March 2023 were included. The obtained data were collected using the questionnaire then entered and analyzed using SPSS version 26.

A total of 101 patients were studied, with the male to female ratio of 1.2:1. Cancer head of pancreas was the commonest malignant cause of obstructive jaundice where as choledocholithiasis was the commonest benign cause. Most of the clinical presentations were abdominal pain 47.6%, yellowish discoloration of the mucous membrane, eyes and skin by 43.6% and itching by 6.9%. Both laboratory investigations and radiological imaging were done which showed elevated liver enzymes 68.3%, low Hb 46.5%, histopathology 62.4%; benign and malignant obstructive jaundice cause by 33.7% and 46.5% respectively. Among all the patients who were surgically managed 29.7% had palliative triple bypass, 14.8% had laparoscopic cholecystectomy and 14.8% had CBD Exploration. The overall complication rate was 50.5%; Death 16.5% surgical site infections 13.9% and long hospital stay 9.9%.

Obstructive jaundice in our setting is more common in males where malignant causes being high in the list

## Introduction

**Obstructive Jaundice** is the yellowish discoloration of the skin, sclera, and mucus membrane due to increased bilirubin concentration in the body fluids as a result of obstruction in the biliary system (Khan, 2019). It is a common problem that occurs when there is an obstruction to the passage of conjugated bile from liver cells to the intestine and is among the common cause of surgical consultation globally (Mabula *et al*., 2014; Dakhore, 2018; Yeola, 2020). The diagnosis is made based on clinical presentations, laboratory and radiological findings. It is among the common causes of surgical consultations globally with an annual incidence of 8-10/100,000 population (Kozarek, 2000; Alrashed *et al*., 2018). In high-incident regions, 242/100,000 cases have been reported annually in Eastern and Central Asia. (Kozarek, 2000; Alrashed *et al*., 2018). An estimated 360 cases of obstructive jaundice were reported at a single tertiary hospital in Ghana (Mercouris *et al*., 2023).

The common **radiological findings** seen in this condition are common bile duct dilatation, cholelithiasis, common bile duct stricture, choledochal cysts, and cancer head of the pancreas as reported in different studies (Dalwani, Shaikh and Devanand, 2013; Gulab Dhar Yadav *et al*., 2022; Khan, 2019). It is among the challenging conditions managed by general surgeons and contributes significantly to high morbidity and mortality despite recent advances both in preoperative diagnosis and postoperative care globally (Mabula *et al*., 2014). The difficulty in diagnosis and treatment of obstructive jaundice is even more pronounced in third-world countries like Tanzania in different hospitals, where the late presentation of the disease coupled with lack of modern diagnostic tools such as CT scans ERCP and MRCP on-surface setbacks (Dalwani, Shaikh and Devanand, 2013; Mabula *et al*., 2014).

It can be caused by many factors both intrahepatic and extrahepatic factors including; benign like cholelithiasis with cholecystitis and malignant conditions like cancer head of pancreas and cholangiocarcinoma and many others which can be identified either preoperatively using radiological diagnostic tools such as sonography, CT scan, MRCP specific for biliary tree scan or postoperatively and ERCP or Intra-operatively(Odongo *et al*., 2022). Early diagnosis and treatment of patients with malignant obstructive jaundice is the speculation of the disease as resection can be done at this stage (Chalya, Kanumba and McHembe, 2011). The most frequent surgical managements offered to obstructive jaundice patients are cholecystectomy, choledochojejunostomy, cholecystojejunostomy, and pancreatoduodenectomy (Gulab Dhar Yadav *et al*., 2022; Shetty *et al*., 2016). Surgeons in resource-limited settings relate the management of obstructive jaundice to difficult hurdles (Odongo *et al*., 2022). When compared to non-jaundiced individuals, surgery in jaundiced patients is associated with a greater risk of postoperative complications(Gulab Dhar Yadav *et al*., 2022; Shetty *et al*., 2016). Treatment outcome of obstructive jaundice are diverse and non-linear among the patients where some patients get either cured without complication; some get severe complications or death (Mabula *et al*., 2014).All these can be explained by different factors such as ; pancreatic malignant, comorbidities, and presentations during the hospital visit (Mulatya, Dharsee and Med, 2022). Understanding the common clinical presentations, etiology and treatment outcome, is important for early detection, decision-making, reduced complications and mortality related to obstructive jaundice in the local community.

## Material and methods

### Study design

This was a cross sectional descriptive, hospital-based study.

### Study area

The study was conducted at General Surgery department, KCMC Hospital-Moshi Municipality. KCMC is a tertiary teaching hospital serving about 15 million people through different departments. The general surgery department receives patients from the local community in the Kilimanjaro region as well as from the nearby regions in the Northern part of Tanzania including Tanga, Arusha, and Manyara, as well as neighboring districts in Kenya near the border. General surgery department receive up to 786 patients per year with different diagnoses. It is well equipped with full time General surgeons specialized in different specialties such as Hepatobiliary, Gastroenterology, Neurosurgery, Cardiothoracic, Pediatric and Plastic and endocrine surgery.

### Study population

All patients who had obstructive jaundice aged 18 years and above who were admitted in General surgery department at KCMC hospital from March 2020 to March 2023.

### Inclusion criteria

All patients aged 18 years and above who were diagnosed with obstructive jaundice and admitted in surgical ward, General Surgery department at KCMC hospital from March 2020 to March 2023.

### Exclusion criteria

Patients who had missing relevant information.

Readmitted patients with the treatment of obstructive jaundice, to prevent data duplications.

### Sampling technique and sample size

Non probability, Convenience sampling was used where all patients aged 18 years and above, who were admitted in surgical ward, General Surgery department at KCMC hospital from March 2020 to March 2023 were included in the study.

### Independent variables

Age, sex, clinicopathological features, Duration of illness, Type of surgery.

### Dependent variables

Early postoperative outcome (surgical site infection, peritonitis, long hospital stays and death)

### Data collection methods and tools

A structured questionnaire was used, with three main parts; first, demographic information including age, sex, level of education, lifestyle, and occupation, second, clinicopathological characteristics including duration of illness, clinical presentation, cause of obstructive jaundice, radiological imaging and laboratory investigations and third, surgical management modalities and early postoperative outcome.

### Data collection procedure

Principal investigator and research assistant identified patient’s registration number from general surgery admission and theater registry books. All files of patients aged 18 years and above from these registry books with a diagnosis of obstructive jaundice from March 2020 to March 2023 were included. The registration number obtained was entered into the EHMS database in order to extract useful information. The information obtained from the EHMS database included; Demographic characteristics, clinicopathological presentations and findings, treatment modalities and outcome. Questionnaire was filled and information entered into SPSS version 26 for analysis. Patient’s index numbers were used during data collection to maintain confidentiality.

### Data management and analysis plans

The collected data was entered and analyzed using SPSS version 26, cleaning was done to ensure data quality and consistency. Mean and standard deviation was used to summarize the numerical data and frequency and proportions was used to summarize categorical variables

### Ethical considerations

Research clearance was obtained from KCMUCo Clinical Research Ethical Review Committee with an Ethical clearance certificate number: **PG 128/2022**.

Formal written consent for patient secondary data extraction was obtained from KCMC hospital administration. The permission to conduct a study was obtained from the head of general surgery department. Patient’s confidentiality was maintained by the use of index numbers and the collected information was only used for the purpose intended by the study. Data were collected from 01^st^ March 2023 to 30^th^ June 2023. Throughout the research activities no access to information that could identify individual participants during or after data collection

### Dissemination of the study findings

The results of this study will be presented to the academic forum of Kilimanjaro Christian Medical University College. Copies of the dissertation will be available in the KCMUCo library, general surgery department and KCMC library and will be submitted to peer review journal for publication.

## Results

### Social demographic characteristics

This study included a total of 101 study participants. The mean (SD) age was 58.7 (16.2) years. However; 35 (34.7%) were aged 61 – 70 years, 53 (52.5%) were males, 44 (43.6%) had the illness for 1 – 3 months, (**Table 1)**;

**Table 1:** Socio demographic characteristics of the study participants (N=101)

| **Characteristics** | **n (%)** |
| --- | --- |
| **Age (mean (SD) years** | 58.7 (16.2) |
| **Age (years)** |  |
| 18 - 30 | 6 (5.9) |
| 31 - 40 | 10 (9.9) |
| 41 - 50 | 15 (14.9) |
| 51 - 60 | 18 (17.8) |
| 61 - 70 | 35 (34.7) |
| > 70 | 17 (16.8) |
| **Sex** |  |
| Female | 48 (47.5) |
| Male | 53 (52.5) |
| **Duration of illness (months)** |  |
| < 1 | 20 (19.8) |
| 1 - 3 | 44 (43.6) |
| 4 - 11 | 20 (19.8) |
| ≥ 12 | 17 (16.8) |

**Table 2:** Causes of obstructive jaundice (N=101)

| Causes | n (%) |
| --- | --- |
| **Benign causes** | **34 (33.7)** |
| Choledocholithiasis | 3 (8.8) |
| Biliary / CBD stricture | 9 (26.5) |
| Chronic pancreatitis | 10 (29.4) |
| Cholelithiasis with cholecystitis | 12 (35.3) |
| **Malignant causes** | **47 (46.5)** |
| Cancer of head of pancreas | 19 (40.4) |
| Cholangiocarcinoma | 4 (8.5) |
| Gall bladder mass/ tumor | 11 (23.4) |
| Periampular mass / tumor | 5 (10.6) |
| Liver hepatoma | 1 (2.1) |
| Duodenal mass | 4 (8.5) |
| Metastatic malignant | 3 (6.4) |
| **Not established** | **20 (19.8)** |

### Clinicopathological presentations

#### Clinical presentation of obstructive jaundice

The clinical presentation among patients with obstructive jaundice includes; 44 (43.6%) had yellowish coloration (eye and skin), 7 (6.9%) had itching / pruritus, 48 (47.6%) had abdominal pain and 2 (1.9%) had others which were pale stool and deep yellow urine, **(Figure 1)**.

**Figure 1:**
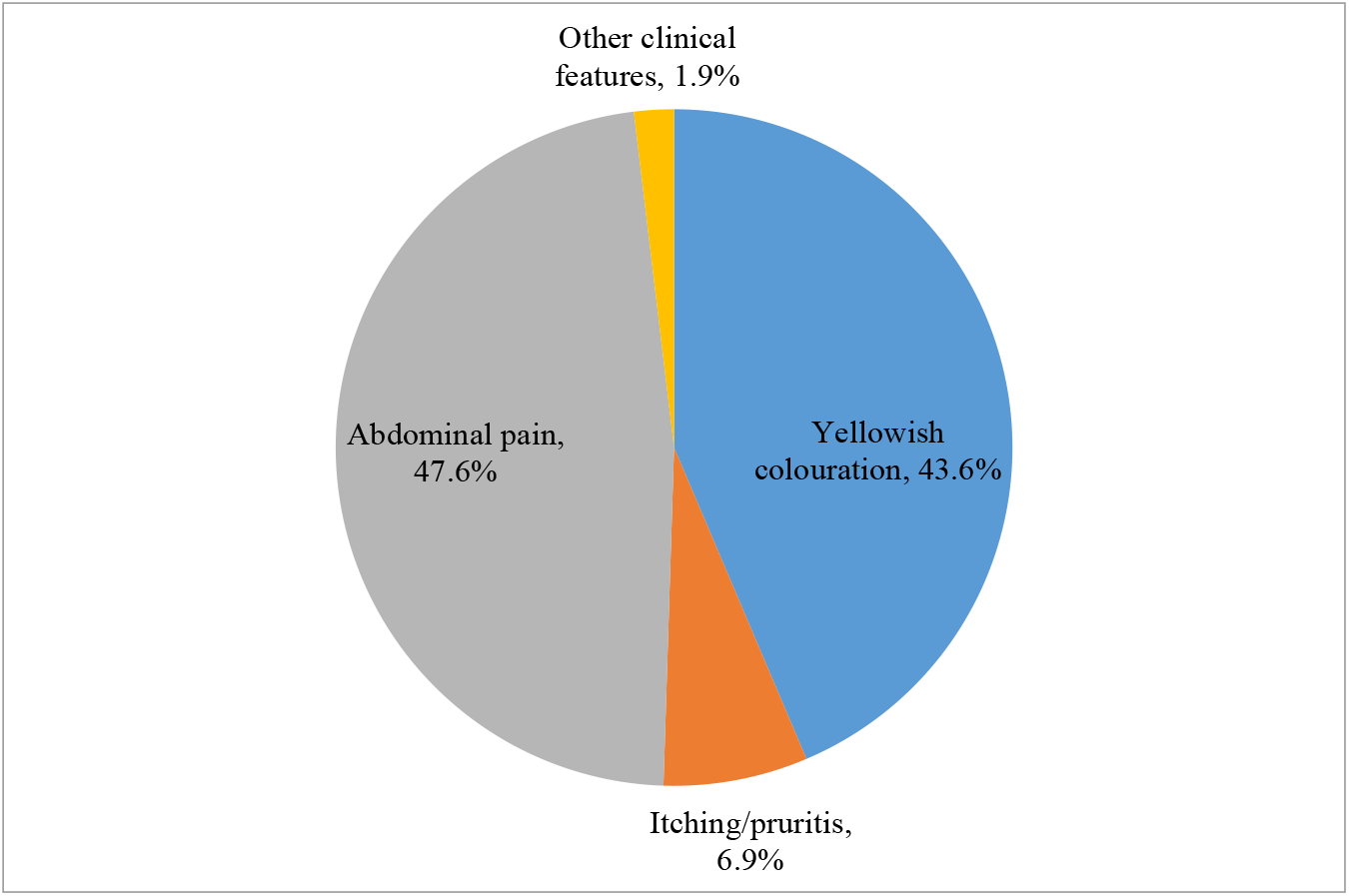
The clinical presentations among patients with obstructive jaundice admitted in surgical ward at KCMC (N=101)

### Causes of obstructive jaundice

Benign causes include; choledocholithiasis, biliary /CBD stricture, chronic pancreatitis and cholelithiasis with cholecystitis. Malignant causes include; Cancer of head of pancreas, cholangiocarcinoma, gallbladder mass/ tumor, periampular mass/ tumor, liver hepatoma, duodenal mass and metastatic malignant, **(Table 4)**.

### Management modalities

The management modalities among obstructive jaundice patients admitted in surgical ward at KCMC include; triple bypass 30(29.7%), open cholecystectomy 10(9.9%), CBD exploration 15(14.8%) and 5(4.9%) were referred to MNH and 17(16.8%) patients were non surgically managed. In the study15 patients had laparoscopic cholecystectomy, these patients presented; Initially with yellowish discoloration of the skin and mucous membrane due to cholelithiasis with cholecystitis, which was medically treated before surgical intervention, **(Figure 2)**.

**Figure 2:**
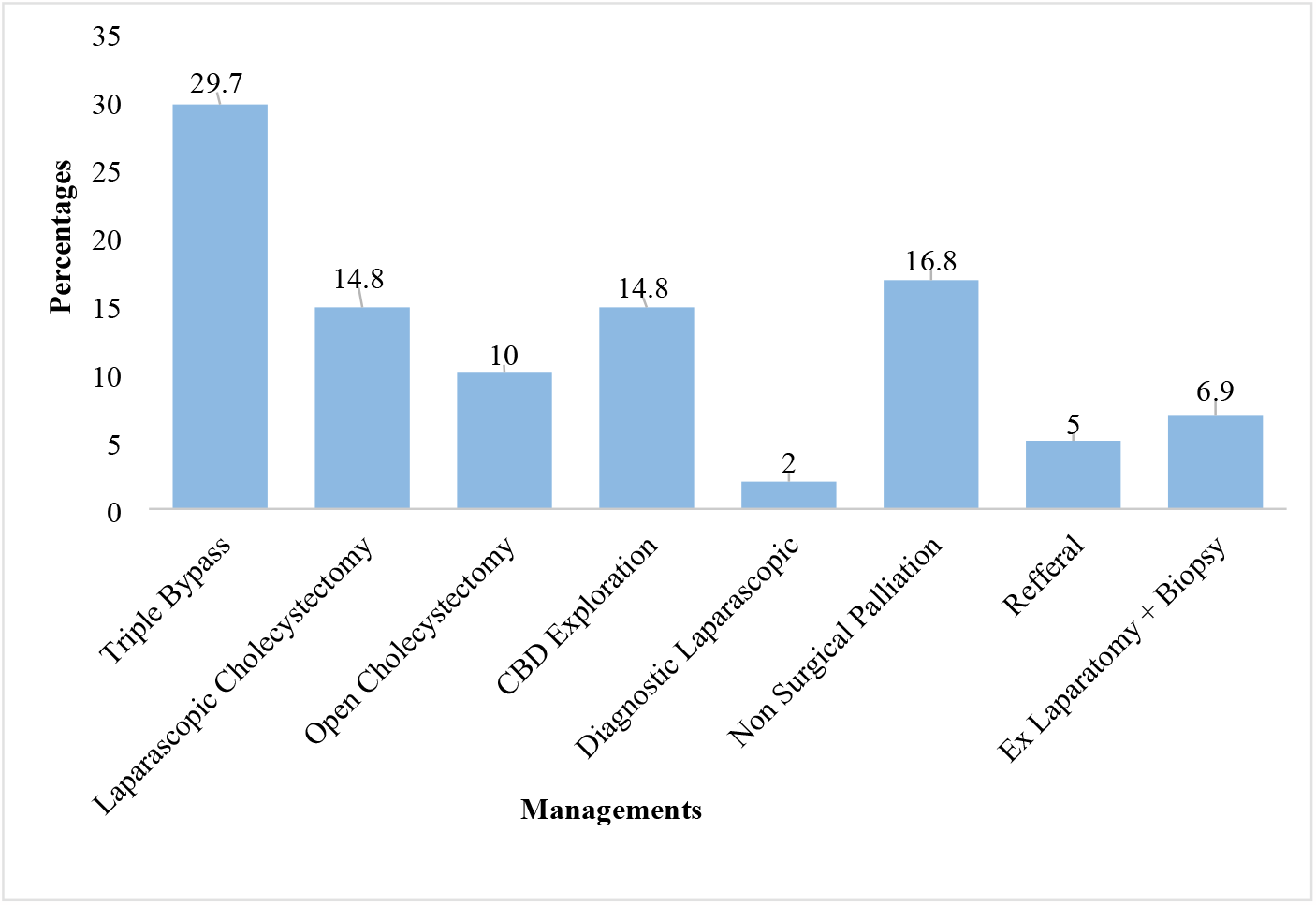
The management modalities among obstructive jaundice patients admitted in surgical ward at KCMC (N=79)

### Postoperative outcome and hospital stay

Among the 79 operated patients 13(16.5%) died, 11(13.9%) developed SSI, 5(6.3%) had peritonitis and 8(10.1%) stayed more than 14 days. Among the 13 post-operative deaths, 12 (92.3%) had malignancy, **(Figure 3 and 4)**.

**Figure 3:**
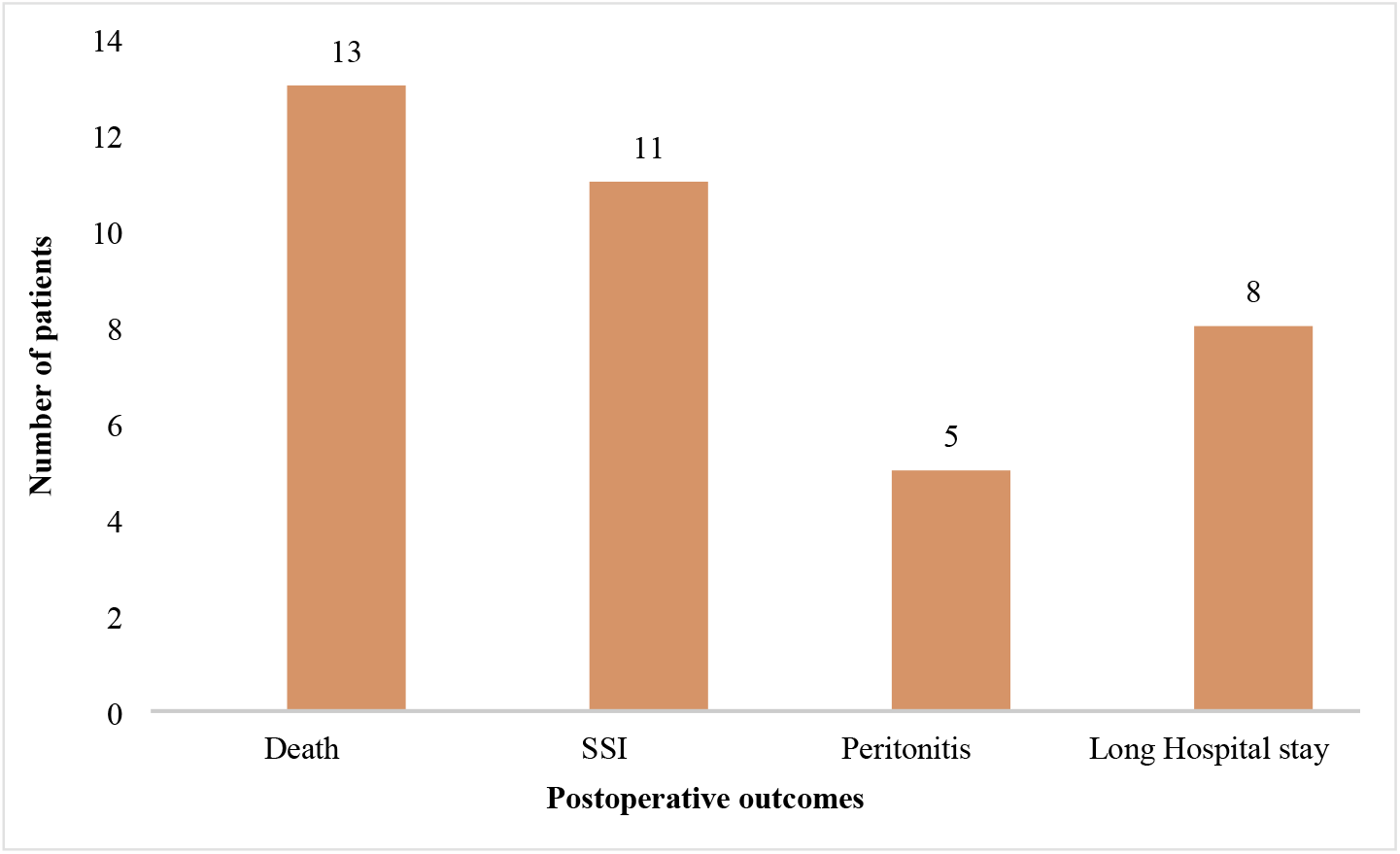
Postoperative outcome among obstructive jaundice patients admitted in surgical ward at KCMC (N=79)

**Figure 4:**
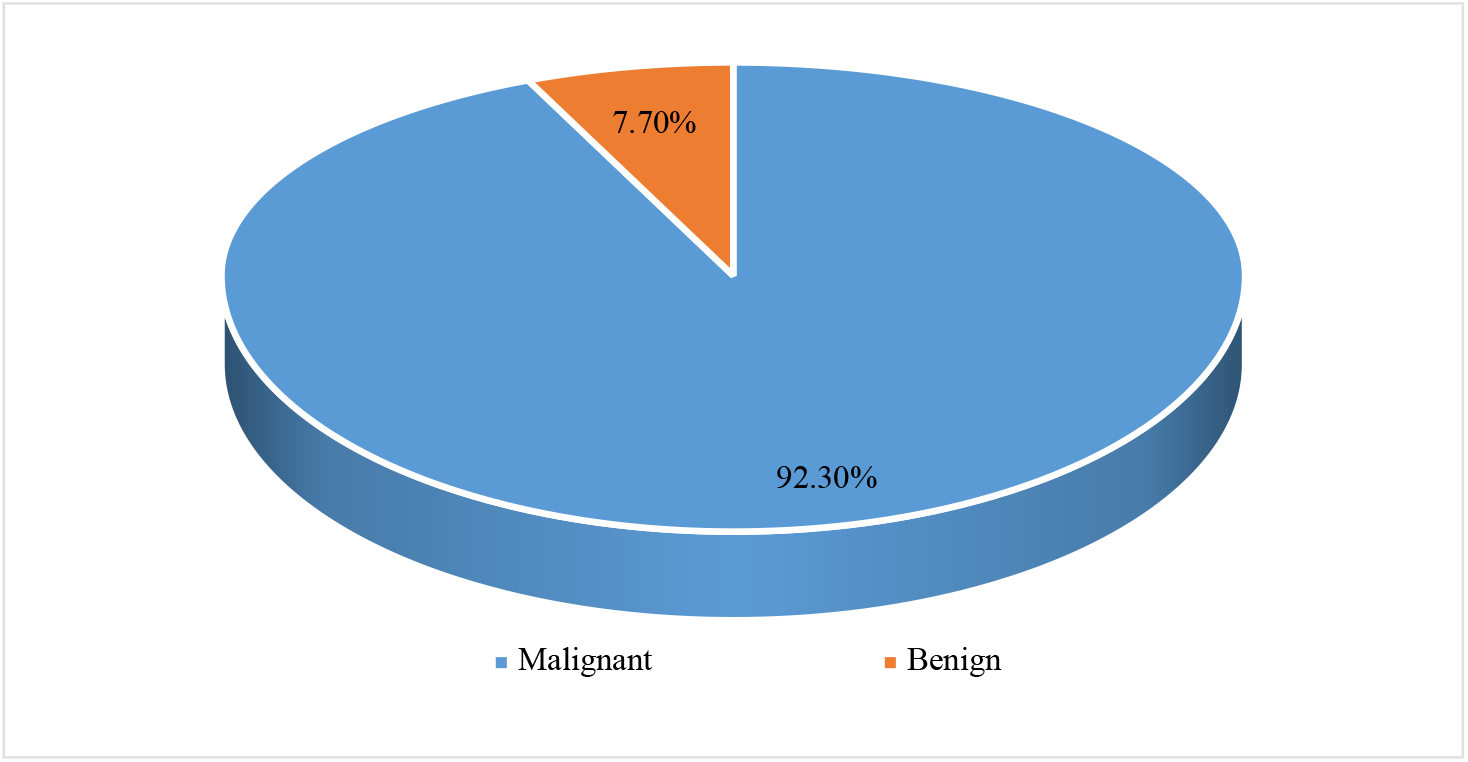
Malignant Vs Benign outcome among obstructive jaundice patients admitted in surgical ward at KCMC (N=13)

## Discussion

The mean age of obstructive jaundice in our study was found to be 58.7 years with prominence of about a quarter of patients at an age of 60-70 years. This has also been reported by other studies, Odongo et al., Gulab et al. Contrary to the evidence of many studies reported female to be more affected by this problem, our results showed more male than female which was 53 (52.5%) as it was reported in the study done in Bugando hospital Mwanza Tanzania by Mabula et al and Chalya et al results.

Minimum duration of illness of these patients in the study was 1-3 months, this was presented by 43.6 % and the rest of patient were ill for longer than this duration; Implying that this is a chronic surgical condition progressively worsening with time. Despite being a chronic surgical condition or illness for a long time their minimum hemoglobin level was > 11 in more than 50% of patients, which looks to be a normal standard, so if this surgical condition could be early diagnosed and planned for early surgical interventions; Can reduce challenges that general surgeon poses in a low resources country as it was reported by the study done in India by Gulab et al

Our study found two cardinal features of presentation which were abdominal pain and yellowish discoloration of the eyes, mucosa and skin. Other manifestation presented in a very low proportions compared to the mentioned cardinal features; these included generalized body itching, deep yellow urine and pale stool. This is in concordance with the study done by Chalya et., al in 2011, which reported 58.65% of yellowish discoloration and 43.1% of body itching. In contrast to the study done in India by Nilesh et., al in 2018, which reported all patients presented with jaundice 100%, but had other features as well including abdominal pain for more than a half of the patients followed by other manifestations like loss of appetite 59.43%, weight loss 57.5%, and itching 41.5%. The study done in India by Gulab et al., 2022 reported Abdominal pain 100% and Icterus 100%, clay-colored stool 58%, fever 54% anorexia 42% and itching, 40% which is inversely to our findings.

All the studied patients did the laboratory investigations relevant to the disease, out of which 61.4% had their histopathology results which revealed malignant causes mainly pancreatic adenocarcinoma, gall bladder adenocarcinoma and invasive adenocarcinoma of pancreas and benign causes mainly cholelithiasis with cholecystitis, other laboratory investigations done were; 53.5% had hemoglobin ≥ 11 g/dl and 68.3% had elevated liver function tests. This is agreeing with the study done in Mwanza by Chalya et., al 2011 and Mabula et., al in 2013 which reported elevated ALAT level in 62.1% and 71.7% respectively. Similar findings were reported by the study done in Uganda which reported elevation of liver enzymes in the majority of the study patients.

Amongst the imaging modalities to find out the cause of obstructive jaundice; Abdominal USS and abdominal CT scan were mostly done, with the following findings; 33.7% had benign cause mainly Cholelithiasis with cholecystitis, 35.3% and 46.5% had malignant causes mainly cancer head of pancreas 40.4%. similar findings were reported in the study done in Mwanza by Chalya et al and Mabula et al which reported cancer head of pancreas being the leading among the malignant causes accounting 64.7%, and 65.1% respectively, the contrary findings were reported among the benign causes where choledocholithiasis was the leading cause by 62.5%) and 51.9% respectively followed by Biliary strictures by 25.0% and 26.9% respectively. In our study very few study patients had MRCP done, there is no clear reason for the low uptake of this diagnostic imaging modality, but it is speculated being because of financial constraints by many patients. This is in contrast to Lorenz et al. who reported the usefulness of MRCP as a special type of diagnostic Imaging modality for hepato-biliary and pancreatic system and can distinguish between malignant and benign causes of obstructive jaundice

The study done in Uganda and Ghana by Odongo et al., 2022 and Mercouris et al., 2023 reported very small percentages of cancer head of pancreas and choledocholithiasis, majority of the causes were gall bladder tumors. The study done in India by Gulab et al., 2022, Belgium by Michael et al., 2019, Bangladesh by Roy et., al 2015 and Pakistan by Siddique et al., 2008, reported high percentages of cancer head of pancreas and choledocholithiasis as the leading cause of obstructive jaundice among the malignant and benign causes respectively.

In our study, Palliative triple bypass has been found to be a more common surgical management offered followed by laparoscopic cholecystectomy and CBD exploration by 29.7%, 14.8% and 14.8% respectively. The study done by Odongo reported similar findings. In contrast, the study done by Chalya and Mabula et., al in Mwanza which reported Cholecystojejunostomy being more offered. Triple bypass procedure technically takes longer time to be performed and may need more stable patients for better outcome. It has a higher risk of mortality and it’s easy to result in disabilities due to post-operative outcome. It is considered to be palliative and this could be a sign of how late patients seek medical attention similarly reported Nilesh et al a study done in India. Our study results show that the mortality rate was 3.3 times higher among the nonoperated patients in comparison to those who were operated. Something which should rise an alarm, on the best to be done to these patients even though presents very late.

Results from the study showed one of the outcome being death (16.5%) of all postoperative patients as a complication. This is a cumulative death which is attributed by different causes including malignant causes (92.3%) of which cancer head of pancreas contributed about 66.7%, followed by cholangiocarcinama25% and gall bladder cancer 8.3%. Malignant causes of deaths dominated in this study and this might be caused by the late symptomatology caused by delayed health seeking behavior of our patients. They present with malignant stages which is incurable by the common surgical procedures and remains to be palliative sort of management. Our results shared the pattern with the results of the study done by Fernandez et al which reported a large proportion of the patients with malignant causes of jaundice reported at the late stages.

## Conclusion

Obstructive jaundice is a chronic condition caused by a variety of preventable and avoidable factors. Early diagnosis and treatment of the causes can prevent further unfavorable outcome and improve patients’ quality of life. This can be achieved by the provision of health education, early seeking of health services as well as building capacity for our health services by having appropriate safe diagnostic and treatment modalities.

## Data Availability

No legal or ethical restriction to data availability

## Recommendations

We recommend early educational intervention especially among school children and other communities. Policymakers should put much effort on improving of the health systems including establishment of affordable and useful medical technology, skills and human resource

We recommend early screening and early diagnosis for timely curative interventions.

We recommend further studies like prospective cohort to be done for the detailed information of the patient.

We recommend building capacity to perform curative surgical interventions for patients with pancreaticobiliary tumors.

## Acknowledgement

First and foremost, thanks be to God for his outworking throughout the process of dissertation writing.

My sincere gratitude to my supervisors Dr. David Msuya (Consultant General Surgeon and Head of General Surgery Department) and Prof. Samwel Chugulu (Consultant Cardiothoracic Surgeon) for their guidance and valuable inputs.

I would also extended my appreciations to Dr Elmes Venant for his availability and an endless support during the period of dissertation writing. Likewise, I would like to give my sincere appreciations to Dr Kennedy Misso, Dr Lele Fabrice and Dr Denis Machaku for their valuable inputs.

Special acknowledgement goes to my spiritual leaders Pastor Frida and Pastor Frank, who have been praying for my success.

Lastly, to my most special and wonderful friends and my beloved children Lionel and Angelprisca for their tireless moral support and great tolerance while i was busy working on my dissertation.

